# Outcomes and Patterns Related to Magnesium in Acute Heart Failure: A Population-Based Study

**DOI:** 10.1101/2025.03.12.25323684

**Authors:** Robert Margaryan, Sunjidatul Islam, Douglas Douver, Finlay A. McAlister, Padma Kaul, Justin A. Ezekowitz

## Abstract

**Importance:** The significance of magnesium as a treatment or prognostic factor is unknown in heart failure despite its frequent use.

**Objective:** To assess the frequency and outcomes of magnesium testing, hypomagnesemia and intravenous (IV) replacement in a large population-based cohort.

**Design, Setting, and Participants:** Retrospective cohort study using linked administrative data from April 2012 - March 2020. Patients with primary diagnosis of HF in the emergency department or hospital were included and the rates and outcomes of magnesium testing, hypomagnesemia and IV replacement were assessed.

**Main Outcome(s) and Measure(s):** The primary clinical outcomes included all-cause and cause specific death and hospitalization. Secondary outcomes included emergency department visits and physicians claims. Other outcomes included factors and rates of serum magnesium testing and hypomagnesemia.

**Results:** Of 78,957 acute heart failure episodes (in 42,763 patients), 58.7% included a serum magnesium measurement. Of the patients who were tested, serum magnesium levels were <0.75 mmol/L in 31.7%, between 0.75 - 0.95 mmol/L in 56.8% and >0.95 mmol/L in 11.5%. Magnesium levels (per 0.02 mmol/L increase) were independently associated with mortality when <0.70 mmol/L [hazard ratio (HR) 0.99 (95% confidence interval (CI) 0.98-0.99); p<0.001] or >0.86 mmol/L [HR 1.04 (95% CI 1.03-1.04); p<0.001]. IV magnesium was given to 13.7% (n=6,333) of those who were tested (29.7% of whom did not have hypomagnesemia); after multivariable adjustment, receiving IV magnesium was associated with a higher short term mortality [HR 1.66 (95% CI 1.4-1.96); p<0.0001] and hospitalization risk [HR 1.36 (95% CI 1.13-1.63); p<0.001].

**Conclusions and Relevance:** Serum magnesium testing is common in patients presenting to the ED or hospital with HF, and low or high magnesium is associated with worse outcomes. Replacement with IV magnesium was associated with worse outcomes even after adjustment, a finding which warrants further study.

## Introduction

Discussions surrounding unnecessary tests and treatments in cardiovascular medicine became more prevalent internationally in 2012 when the Choosing Wisely campaign was launched.^1^ Decisions about whether to administer a test or provide a therapy are even more involved in complex conditions like heart failure (HF) which is associated with frequent emergency department visits, hospital admissions, and a 5-year survival rate of roughly 50%.^2^ While many tests, treatments and procedures are being evaluated closely to determine their necessity, to our knowledge the frequency and patterns of serum magnesium level testing and intravenous (IV) magnesium treatments have not been evaluated.

Electrolyte imbalances such as hypomagnesemia are frequently observed in HF patients with 19-53% having hypomagnesemia.^3–7^ However, there is a limited understanding of the involvement of magnesium in the pathophysiology of HF particularly in the acute setting. Several studies have shown treatment with magnesium to be associated with greater conductive and vascular stability in both patients with acute and chronic HF.^3–9^ Earlier studies in this area also indicated that there may be an association between hypomagnesemia and increased mortality risk in patients with HF.^3^ However, results have varied, and consequently there is no consensus on whether hypomagnesemia is associated with adverse outcomes in patients with acute or chronic HF.^5, 10–11^ Further, while large clinical trials have shown that IV magnesium does not improve survival in acute myocardial infarction, it’s efficacy as a treatment in acute heart failure has not been evaluated.^12–13^

Thus, to address these knowledge gaps, we designed this study to explore the frequency and factors related to magnesium testing and treatment, as well as any associations between hypomagnesemia and IV magnesium treatments with patient outcomes in patients with acute heart failure (AHF).

## Methods

This is a retrospective cohort study which used the discharge abstract database (DAD) and the national ambulatory care reporting system (NACRS) data from April 1, 2012 to March 31, 2020 in Alberta, Canada. This period of time was chosen to avoid overlap with the abnormal care conditions stemming from the COVID-19 pandemic. Details of other sources of data are available in the **Supplemental Material**. All of the diagnoses were complied with International Classification of Diseases-10th Revision (ICD-10) codes (**Supplemental Table 1**) and this study is reported in accordance with Strengthening the Reporting of Observational Studies in Epidemiology (STROBE) guidelines.

### Patient Population

Patients included in the cohort were adults (> 18 years old) who visited the emergency department or were hospitalized with a primary diagnosis of HF (ICD-10: I50.x) between April 01, 2012 and March 31, 2020. Recognizing the discontinuous nature of IV magnesium replacement in patients with HF, we analyzed each hospitalization or ED visit (except those within 48 hours) as discrete episodes of care and consequently analyses were done at the episode level rather than the patient level.

### Serum Magnesium Definition

Serum magnesium tests and other laboratory tests were captured from the laboratory database if the test was done within ± 1 day of HF episodes. The lowest values of serum magnesium concentration were chosen when multiple tests were done during a single episode as they were the most representative values. For other tests, the tests closest to the episode start date were selected for the analysis. The main analysis evaluated serum magnesium as a continuous variable. Serum magnesium levels were considered to be indicative of hypomagnesemia if they were lower than 0.75 mmol/L, however a range of cutoffs were explored.^10, 14–15^

### Intravenous Magnesium Therapy Classification

Patient episodes were considered in the analysis when the magnesium compound administered was formulated as magnesium sulfate (MgSO_4_) as this is the only available IV magnesium formulation in Canada.

### Outcomes

The primary clinical outcomes of interest included all-cause death and hospitalization (all-cause, CV related and HF-related). Secondary outcomes of interest included ED visits (all-cause, CV related and HF-related) and Physicians claims (all-cause, CV related and HF-related). Other outcomes included factors and rates of serum magnesium testing and hypomagnesemia. The time to event was measured as the time from each episode to the first outcome event that occurred during the follow-up.

### Statistical Analysis

Categorical variables were summarized as counts with proportions, and compared between/across groups using Chi-square tests. Continuous variables were summarized as medians with interquartile range (IQR). Mann-Whitney U test was used to compare continuous variables between two groups, and the Kruskal-Wallis test was applied when comparing more than two groups.

We used multivariable binary logistic regression models with generalized estimating equation (GEE) to determine (i) the factors associated with magnesium testing and (ii) the factors associated with hypomagnesemia. We included a number of variables such as patient demographics, comorbidities, and medications (see **Supplemental Material** for full list).

The crude rate of outcomes in groups receiving versus not receiving magnesium therapy was presented as the rate per 100 person-months. Time-varying Cox regression models with weighting were used to determine the change in risk of mortality and other clinical outcomes with IV magnesium sulfate administration. We limited this analysis to patients aged 60 to 90 years old, with a maximum of three HF episodes, due to the lower number of IV magnesium administration events among patients who did not meet the above criteria preventing effective group balancing. Outcomes were reported as weighted hazard ratios (HR) with 95% confidence interval (CI). To get the propensity of receiving magnesium supplementation at each episode, we constructed a multivariable binary logistic regression model including a number of variables (see **Supplemental Material** for full list). After examining the interaction between variables, an interaction term for age and serum magnesium value was included in the propensity model. The propensity score was then calculated and included as a time varying covariate and the balance of variables between groups was assessed using the standardized mean differences; variables were considered well balanced if the standardized mean differences ranged from -0.2 to 0.2. Overlap weighting was used instead of inverse probability weighting to avoid the influence of observations with extreme weight on the outcomes when balancing the covariates.^16^ The overlap weighting approach assigned each patient a weight proportional to his or her probability of being in the opposite treatment group, and as a result we observed an exact balance of covariates between the treatment groups (**Supplemental Figure 1**). Further, a negation endpoint was used to further explore the potential presence of bias in the time-varying Cox regression model. Further to this, the association between IV magnesium and urinary tract infections (UTIs) as well as hip fractures was evaluated. Both conditions are associated with increased frailty and poorer overall health unrelated to magnesium pathophysiology or the cardiovascular system.

Two sided p-values less than 0.05 were considered statistically significant. All analyses were done with SAS 9.4 version (SAS Institute Inc).

This study was approved by the University of Alberta Research Ethics Board (Pro00010852). Individual patient consent was not deemed necessary by the board given that Alberta Health Services provided deidentified administrative data.

## Results

Our study included 78,957 episodes of hospitalization or ED visits for HF (in 42,763 patients) followed for a median of 1.9 years [Interquartile range (IQR) 0-3.8] after their first episode of care), and in 58.7% of episodes, serum magnesium was measured. The median age of the patient cohort was 80 years (IQR 70, 87) with 47.2% of patients being female. Among 46,363 episodes during which magnesium were measured, patients had serum magnesium level <0.75 mmol/L in 31.7% of episodes, between 0.75 - 0.95 mmol/L in 56.8% of episodes and >0.95 mmol/L 11.5% of episodes. Serum magnesium level tests were more likely to occur in episodes at a tertiary hospital (23.7% vs 5.7%; p<0.0001), for patients that were admitted rather than discharged from the ED (69.1% vs 42.3%; p<0.0001) and with a longer length of stay (5 (IQR 1,11) vs 1 (IQR 1,5) days; p<.0001) (Table 1). Patients had higher BNP (858 (IQR 478, 1580) vs 746 (IQR 413, 1364); p<.0001) and NT-proBNP (1908 (IQR 696, 5528) vs 1420 (IQR 531, 3877); p<.0001) values in episodes with a magnesium test compared with episodes without a magnesium test (Table 1). Tested episodes also had higher Charlson Comorbidity Index (5 (IQR 3, 7) vs 4 (IQR 2, 6); p<0.0001) **(Table 1)**. However, there was no difference in potassium levels, medication use, or ejection fraction between the episodes of the two groups **(Supplemental Figure 2)**.

**Table 1.** Characteristics of patients in the cohort according to presence or absence of magnesium test during hospitalization. Total number of hemoglobin tests; 75,953 (96.2%). Total number of serum creatinine tests; 76,262 (96.6%). Total number of sodium tests; 50,305 (63.7%). Total number of potassium tests; 50,403 (63.8%). Abbreviations: IQR, interquartile range; ED, emergency department; HFmrEF, heart failure with mildly reduced ejection fraction; HFrEF, heart failure with reduced ejection fraction; HFpEF, heart failure with preserved ejection fraction; CAD, coronary artery disease; PVD, peripheral vascular disease; TIA, transient ischemic attack; COPD, chronic obstructive pulmonary disease; ACEI, angiotensin converting enzyme inhibitor; ARB, angiotensin-receptor blocker; ARNI, angiotensin receptor neprilysin inhibitor; MRA, mineralocorticoid receptor antagonist; LVEF, left ventricular ejection fraction; BNP, B-type natriuretic peptide; NTproBNP, N-type proBNP

| Variable Label | Statistic | Episodes with no tests | Episodes with tests | p-value |
| --- | --- | --- | --- | --- |
| Total N | N | 32,594 | 46,363 |  |
| Age (years) | Median (IQR) | 81 (72, 88) | 80 (69, 87) | <.0001 |
| Female Sex | n (%) | 15,609 (47.9) | 21,620 (46.6) | 0.0005 |
| Residence |  |  |  |  |
| Rural | n (%) | 10,692 (32.8) | 7,938 (17.1) | <.0001 |
| Urban | n (%) | 21,902 (67.2) | 38,425 (82.9) |  |
| Tertiary hospital | n (%) | 1,860 (5.7) | 10,968 (23.7) | <.0001 |
| Clinical setting |  |  |  |  |
| Discharged from ED | n (%) | 18,799 (57.7) | 14,332 (30.9) | <.0001 |
| Admitted | n (%) | 13,795 (42.3) | 32,031 (69.1) |  |
| Length of episode | Median (IQR) | 1 (1, 5) | 5 (1, 11) | <.0001 |
| Fiscal year |  |  |  |  |
| 2012 | n (%) | 4,033 (12.4) | 4,955 (10.7) | <.0001 |
| 2013 | n (%) | 3,911 (12.0) | 5,457 (11.8) |  |
| 2014 | n (%) | 4,166 (12.8) | 5,855 (12.6) |  |
| 2015 | n (%) | 4,070 (12.5) | 5,981 (12.9) |  |
| 2016 | n (%) | 4,147 (12.7) | 6,351 (13.7) |  |
| 2017 | n (%) | 4,388 (13.5) | 6,616 (14.3) |  |
| 2018 | n (%) | 4,566 (14.0) | 6,858 (14.8) |  |
| 2019 | n (%) | 3,313 (10.2) | 4,290 (9.3) |  |
| HF subtype |  |  |  |  |
| HFmrEF | n (%) | 1,002 (3.1) | 1,745 (3.8) | <.0001 |
| HFpEF | n (%) | 4,549 (14.0) | 6,713 (14.5) |  |
| HFrEF | n (%) | 1,876 (5.8) | 3,910 (8.4) |  |
| Missing | n (%) | 25,167 (77.2) | 33,995 (73.3) |  |
| **Comorbidities** |  |  |  |  |
| Diabetes | n (%) | 14,175 (43.5) | 21,662 (46.7) | <.0001 |
| Hypertension | n (%) | 28,516 (87.5) | 40,372 (87.1) | 0.0887 |
| Dyslipidemia | n (%) | 5,798 (17.8) | 9,125 (19.7) | <.0001 |
| CAD | n (%) | 17,153 (52.6) | 24,709 (53.3) | 0.0639 |
| PVD | n (%) | 5,451 (16.7) | 7,429 (16.0) | 0.0087 |
| Atrial Fibrillation | n (%) | 16,508 (50.6) | 24,688 (53.2) | <.0001 |
| Thromboembolism | n (%) | 4,940 (15.2) | 7,052 (15.2) | 0.8344 |
| Asthma | n (%) | 3,931 (12.1) | 5,240 (11.3) | 0.0011 |
| COPD | n (%) | 12,718 (39.0) | 17,481 (37.7) | 0.0002 |
| Cancer | n (%) | 5,671 (17.4) | 7,749 (16.7) | 0.0116 |
| Sleep Apnea | n (%) | 3,079 (9.4) | 4,965 (10.7) | <.0001 |
| Depression | n (%) | 6,111 (18.7) | 9,365 (20.2) | <.0001 |
| Dementia | n (%) | 3,386 (10.4) | 4,875 (10.5) | 0.5677 |
| Charlson Comorbidity Index | Median (IQR) | 4 (2, 6) | 5 (3, 7) | <.0001 |
| **Medications** |  |  |  |  |
| ACEi | n (%) | 17,328 (53.2) | 24,400 (52.6) | 0.1382 |
| ARB | n (%) | 10,815 (33.2) | 15,136 (32.6) | 0.1156 |
| ARNi | n (%) | 403 (1.2) | 529 (1.1) | 0.2216 |
| MRA | n (%) | 9,320 (28.6) | 12,327 (26.6) | <.0001 |
| ACEi/ARB/ARNi | n (%) | 25,385 (77.9) | 35,541 (76.7) | <.0001 |
| Beta blocker | n (%) | 23,250 (71.3) | 33,320 (71.9) | 0.1002 |
| Digoxin | n (%) | 4,426 (13.6) | 5,995 (12.9) | 0.0080 |
| Statin | n (%) | 18,681 (57.3) | 27,237 (58.7) | <.0001 |
| **Imaging** |  |  |  |  |
| LVEF test | n (%) | 7,427 (22.8) | 12,368 (26.7) | <.0001 |
| LVEF values | Median (IQR) | 55 (40, 65) | 53 (35, 65) | <.0001 |
| **Laboratory** |  |  |  |  |
| Hemoglobin | n (%) | 29,734 (91.2) | 46,219 (99.7) | <.0001 |
| Hemoglobin, g/dl | Median (IQR) | 12.1 (10.6, 13.5) | 11.9 (10.3-13.4) | <.0001 |
| Serum Creatinine | n (%) | 29,989 (92.0) | 46,273 (99.8) | <.0001 |
| Serum creatinine value | Median (IQR) | 106.0 (81.0, 146.0) | 113.0 (85.0, 161.0) | <.0001 |
| Sodium | n (%) | 17,839 (54.7) | 32,466 (70.0) | <.0001 |
| Sodium value | Median (IQR) | 139.0 (136.0, 142.0) | 138.0 (135.0, 141.0) | <.0001 |
| Potassium | n (%) | 17,894 (54.9) | 32,509 (70.1) | <.0001 |
| Potassium value | Median (IQR) | 4.0 (3.8, 4.3) | 4.0 (3.0, 4.2) | <.0001 |
| BNP | n (%) | 4,067 (12.5) | 17,813 (38.4) | <.0001 |
| BNP value | Median (IQR) | 746.0 (413.0, 1364.0) | 858.0 (478.0, 1580.0) | <.0001 |
| NTproBNP | n (%) | 18,068 (55.4) | 20,748 (44.8) | <.0001 |
| NTproBNP value | Median (IQR) | 1420.0 (531.0, 3877.0) | 1908.0 (696.0, 5528.0) | <.0001 |

Magnesium levels (per 0.02 mmol/L increase) were independently associated with mortality when they were less than 0.70 mmol/L [hazard ratio (HR) 0.99 (95% confidence interval (CI) 0.98-0.99); p<0.001] or greater than 0.86 mmol/L [HR 1.04 (95% CI 1.03-1.04); p<0.001] **(Figure 1)**. The association between hypomagnesemia and various variables were evaluated at various serum magnesium cutoffs, results are presented for the cut off of 0.75 mmol/L **(Supplemental Figures 3,4,5)**. Episodes with hypomagnesemia were associated with higher rates of admission to the hospital compared with normal magnesium levels and hypermagnesemia respectively (72.4% vs 67.8% vs 66.3%; p<0.0001) **(Table 2)**. Patients in episodes with hypomagnesemia also had longer durations of stay (6 (IQR 1, 13) vs 5 (IQR 1, 10) vs 4 (1,10); p<0.0001) **(Table 2)**.

**Figure 1.**
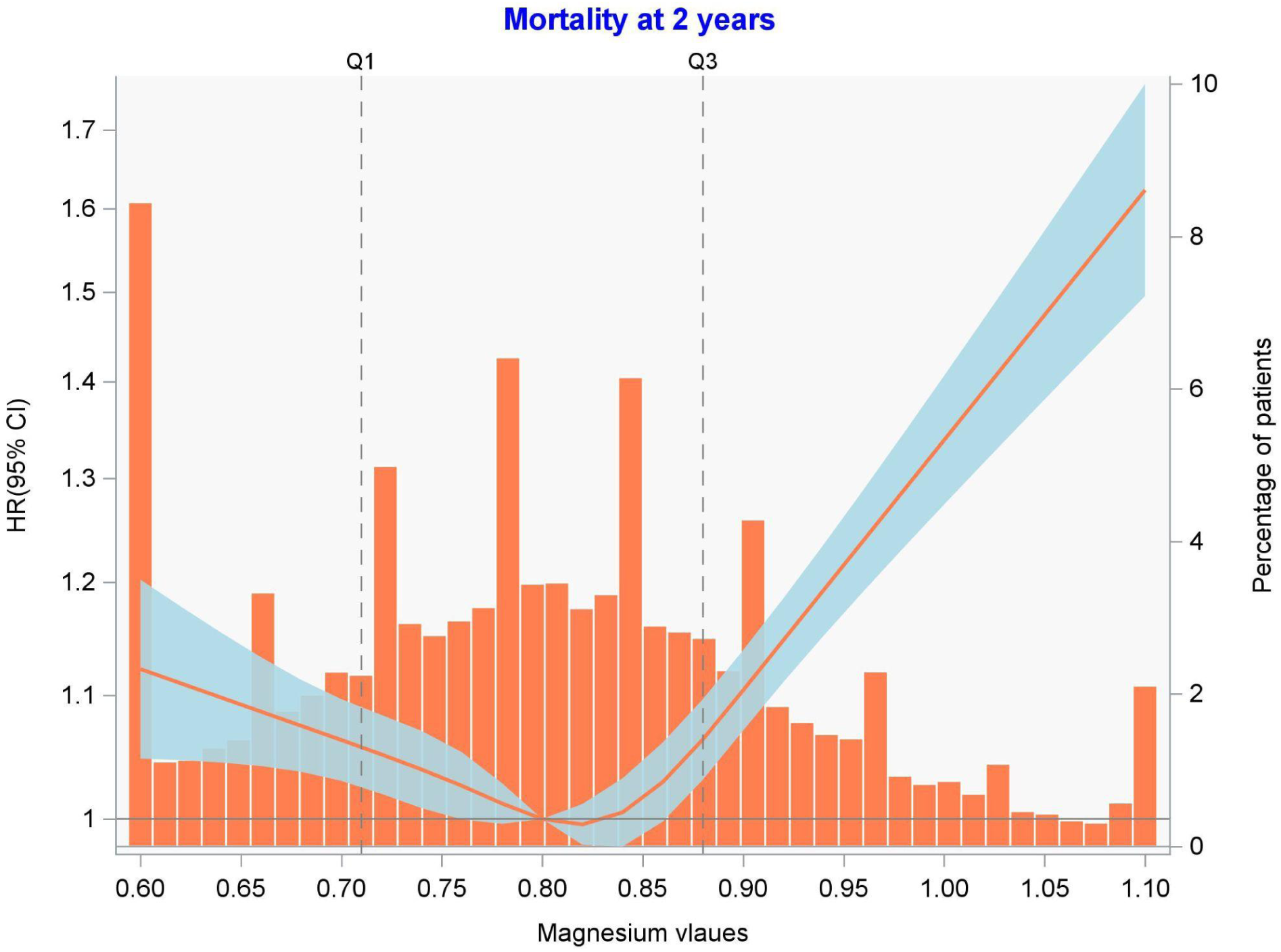
Patient magnesium concentration distribution with a hazard ratio curve for mortality at 2 years overlayed. Abbreviations: HR, hazard ratio; CI, confidence interval.

**Table 2.** Characteristics of patients in the cohort with hypomagnesemia (<0.75 mmol/L), normal magnesemia (0.75-0.95 mmol/L), and hypermagnesemia (>0.95 mmol/L). Total number of hemoglobin tests; 46,219 (99.7%). Total number of serum creatinine tests; 46,273 (99.8%). Total number of sodium tests; 32,466 (70.0%). Total number of potassium tests; 32,509 (70.1%). Abbreviations: IQR, interquartile range; ED, emergency department; HFmrEF, heart failure with mildly reduced ejection fraction; HFrEF, heart failure with reduced ejection fraction; HFpEF, heart failure with preserved ejection fraction; CAD, coronary artery disease; PVD, peripheral vascular disease; TIA, transient ischemic attack; COPD, chronic obstructive pulmonary disease; ACEI, angiotensin converting enzyme inhibitor; ARB, angiotensin-receptor blocker; ARNI, angiotensin receptor neprilysin inhibitor; MRA, mineralocorticoid receptor antagonist; LVEF, left ventricular ejection fraction; BNP, B-type natriuretic peptide; NTproBNP, N-type proBNP

| Variable Label | Statistic | Magnesium<br><0.75 mmol/L | Magnesium<br>0.75-0.95<br>mmol/L | Magnesium<br>>0.95 mmol | p-value |
| --- | --- | --- | --- | --- | --- |
| Total N | N | 14,699 | 26,323 | 5,341 |  |
| Age | Median<br>(IQR) | 77 (68, 85) | 80 (70, 87) | 82 (73, 88) | <.0001 |
| Female Sex | n (%) | 7,365 (50.1) | 11,909 (45.2) | 2,346 (43.9) | <.0001 |
| Residence |  |  |  |  |  |
| Rural | n (%) | 2,710 (18.4) | 4,360 (16.6) | 868 (16.3) | <.0001 |
| Urban | n (%) | 11,989 (81.6) | 21,963 (83.4) | 4,473 (83.7) |  |
| Tertiary hospital | n (%) | 3,676 (25.0) | 6,122 (23.3) | 1,170 (21.9) | <.0001 |
| Clinical setting |  |  |  |  |  |
| Discharged from ED | n (%) | 4,053 (27.6) | 8,481 (32.2) | 1,798 (33.7) | <.0001 |
| Admitted | n (%) | 10,646 (72.4) | 17,842 (67.8) | 3,543 (66.3) |  |
| Length of episode | Median<br>(IQR) | 6 (1, 13) | 5 (1, 10) | 4 (1, 10) | <.0001 |
| Fiscal year |  |  |  |  |  |
| 2012 | n (%) | 1,522 (10.4) | 2,846 (10.8) | 587 (11.0) | <.0001 |
| 2013 | n (%) | 1,712 (11.6) | 3,127 (11.9) | 618 (11.6) |  |
| 2014 | n (%) | 2,035 (13.8) | 3,225 (12.3) | 595 (11.1) |  |
| 2015 | n (%) | 1,951 (13.3) | 3,464 (13.2) | 566 (10.6) |  |
| 2016 | n (%) | 2,028 (13.8) | 3,591 (13.6) | 732 (13.7) |  |
| 2017 | n (%) | 2,070 (14.1) | 3,755 (14.3) | 791 (14.8) |  |
| 2018 | n (%) | 2,169 (14.8) | 3,844 (14.6) | 845 (15.8) |  |
| 2019 | n (%) | 1,212 (8.2) | 2,471 (9.4) | 607 (11.4) |  |
| HF type |  |  |  |  |  |
| HFmEF | n (%) | 638 (4.3) | 953 (3.6) | 154 (2.9) | <.0001 |
| HFpEF | n (%) | 2,332 (15.9) | 3,746 (14.2) | 635 (11.9) |  |
| HFrEF | n (%) | 1,317 (9.0) | 2,268 (8.6) | 325 (6.1) |  |
| Missing | n (%) | 10,412 (70.8) | 19,356 (73.5) | 4,227 (79.1) |  |
| **Comorbidities** |  |  |  |  |  |
| Diabetes | n (%) | 8,457 (57.5) | 10,782 (41.0) | 2,423 (45.4) | <.0001 |
| Hypertension | n (%) | 13,030 (88.6) | 22,547 (85.7) | 4,795 (89.8) | <.0001 |
| Hyperlipidemia | n (%) | 2,992 (20.4) | 5,024 (19.1) | 1,109 (20.8) | 0.0009 |
| CAD | n (%) | 7,816 (53.2) | 13,922 (52.9) | 2,971 (55.6) | 0.0012 |
| PVD | n (%) | 2,412 (16.4) | 4,047 (15.4) | 970 (18.2) | <.0001 |
| Atrial Fibrillation | n (%) | 7,640 (52.0) | 14,122 (53.6) | 2,926 (54.8) | 0.0003 |
| Cerebrovascular-Stroke/TIA | n (%) | 18 (0.1) | 31 (0.1) | 4 (0.1) | 0.6571 |
| Thromboembolism | n (%) | 2,395 (16.3) | 3,846 (14.6) | 811 (15.2) | <.0001 |
| Asthma | n (%) | 1,710 (11.6) | 2,936 (11.2) | 594 (11.1) | 0.3071 |
| COPD | n (%) | 6,000 (40.8) | 9,585 (36.4) | 1,896 (35.5) | <.0001 |
| Anemia | n (%) | 4,768 (32.4) | 7,504 (28.5) | 1,858 (34.8) | <.0001 |
| Sleep Apnea | n (%) | 1,782 (12.1) | 2,632 (10.0) | 551 (10.3) | <.0001 |
| Depression | n (%) | 3,244 (22.1) | 5,171 (19.6) | 950 (17.8) | <.0001 |
| Dementia | n (%) | 1,465 (10.0) | 2,844 (10.8) | 566 (10.6) | 0.0291 |
| Charlson Comorbidity Index | Median (IQR) | 5 (3, 7) | 4 (2, 6) | 5 (3, 7) | <.0001 |
| **Medications** |  |  |  |  |  |
| ACEi | n (%) | 7,980 (54.3) | 13,562 (51.5) | 2,858 (53.5) | <.0001 |
| ARB | n (%) | 4,822 (32.8) | 8,372 (31.8) | 1,942 (36.4) | <.0001 |
| ARNi | n (%) | 154 (1.0) | 312 (1.2) | 63 (1.2) | 0.4355 |
| MRA | n (%) | 3,689 (25.1) | 6,863 (26.1) | 1,775 (33.2) | <.0001 |
| ACEi/ARB/ARNi | n (%) | 11,588 (78.8) | 19,725 (74.9) | 4,228 (79.2) | <.0001 |
| Beta blocker | n (%) | 10,678 (72.6) | 18,585 (70.6) | 4,057 (76.0) | <.0001 |
| Digoxin | n (%) | 1,788 (12.2) | 3,390 (12.9) | 817 (15.3) | <.0001 |
| Statin | n (%) | 9,178 (62.4) | 14,923 (56.7) | 3,136 (58.7) | <.0001 |
| **Imaging** |  |  |  |  |  |
| LVEF test | n (%) | 4,287 (29.2) | 6,967 (26.5) | 1,114 (20.9) | <.0001 |
| LVEF | Median (IQR) | 53 (35, 65) | 53 (34, 65) | 53 (36, 65) | 0.0335 |
| **Laboratory tests** |  |  |  |  |  |
| Hemoglobin test | n (%) | 14,651 (99.7) | 26,244 (99.7) | 5,324 (99.7) | 0.8939 |
| Hemoglobin (g/dl) | Median (IQR) | 11.8 (10.3, 13.3) | 12.1 (10.5, 13.6) | 11.3 (9.7, 12.9) | <.0001 |
| Serum creatinine | n (%) | 14,671 (99.8) | 26,267 (99.8) | 5,335 (99.9) | 0.3127 |
| Serum creatinine value | Median (IQR) | 103.0 (79.0, 142.0) | 111.0 (85.0, 155.0) | 169.0 (122.0, 251.0) | <.0001 |
| Sodium | n (%) | 10,531 (71.6) | 18,218 (69.2) | 3,717 (69.6) | <.0001 |
| Sodium value | Median (IQR) | 138.0 (135.0, 141.0) | 139.0 (136.0, 141.0) | 138.0 (135.0, 141.0) | <.0001 |
| Potassium | n (%) | 10,542 (71.7) | 18,239 (69.3) | 3,728 (69.8) | <.0001 |
| Potassium value | Median (IQR) | 4.0 (3.0, 4.0) | 4.0 (3.2, 4.2) | 4.0 (4.0, 4.8) | <.0001 |
| BNP | n (%) | 5,562 (37.8) | 10,365 (39.4) | 1,886 (35.3) | <.0001 |
| BNP value | Median (IQR) | 544.6 (198.4, 1220.0) | 572.8 (214.0, 1237.6) | 708.1 (238.1, 1679.7) | <.0001 |
| NTproBNP | n (%) | 6,626 (45.1) | 11,683 (44.4) | 2,439 (45.7) | 0.1435 |
| NTproBNP value | Median (IQR) | 1830.0 (676.0, 5305.0) | 1898.0 (696.0, 5385.0) | 2209.0 (781.0, 7421.0) | <.0001 |

IV magnesium was given in 13.7% (n=6,333) of episodes which featured a serum magnesium test, and in 30.3% (n=4,451) of episodes with hypomagnesemia. Among patient episodes which received IV Mg (n=6,333), 70.3% (n=4,451) of episodes had magnesium levels <0.75 mmol/L, 27.5% (n=1,744) had magnesium levels 0.75-0.95 mmol/L and 2.2% (n=138) had magnesium levels above 0.95 mmol/L. Patients who had a test had similar rates of death (1539 deaths at a rate of 2.7 per 100 person months vs 9,010 deaths at a rate of 2.6 per 100 person months) and hospitalizations for any cause (2176 at a rate of 5.8 per 100 person months vs 13,339 at a rate of 5.8 per 100 person months) regardless of whether magnesium was administered or not **(Supplemental Table 2)**. Having an urban residence [aOR 1.55 (95% CI 1.42-1.69); p<0.001], visiting a tertiary hospital [aOR 1.35 (95% CI 1.26-1.45); p<0.001], and being admitted to the hospital [aOR 2.41 (95% CI 2.22-2.62); p<0.001] were associated with an increased likelihood of receiving IV magnesium **(Supplemental Table 3)**. Similarly, patients with coronary artery disease [aOR 1.13 (95% CI 1.06-1.21); p<0.001] and atrial fibrillation [aOR 1.17 (95% CI 1.09-1.25); p<0.001] were also more likely to receive IV magnesium **(Supplemental Table 3)**. Additionally, every 0.1 mmol/L reduction in serum magnesium concentration from 0.8 mmol/L was associated with a 308% increase in likelihood of receiving IV magnesium [aOR 4.08 (95% CI 3.92-4.25); p<0.001] **(Supplemental Table 3)**.

After weighting, receiving IV magnesium was associated with a higher 7 day mortality risk [HR 1.66 (95% CI 1.4-1.96); p<0.0001], that persisted from 7-30 days [HR 1.47 (95% CI 1.12-1.93); p=0.005] **(Figure 2)**. In the 7 days following the infusion of magnesium a pattern of mortality risk according to serum magnesium concentration was evident as patients with normal magnesium levels had higher risk of death compared to both patients with hypomagnesemia and patients with hypermagnesemia **(Figure 3)**. IV magnesium was also associated with a higher risk of 7 day [HR 1.36 (95% CI 1.13-1.63); p<0.001] all-cause hospitalization and a reduction in the risk of 7 day all-cause ED visits [HR 0.63 (95% CI 0.50-0.78); p<0.001] after weighting **(Figure 2)**. IV magnesium was associated with an increase in risk of 30-60 day CV physician claims after weighting [HR 1.26 (95% 1.06-1.51); p=0.0093] **(Figure 2)**. All patient outcomes at all other time points were similar regardless of IV magnesium administration **(Supplemental Figure 6)**.

**Figure 2.**
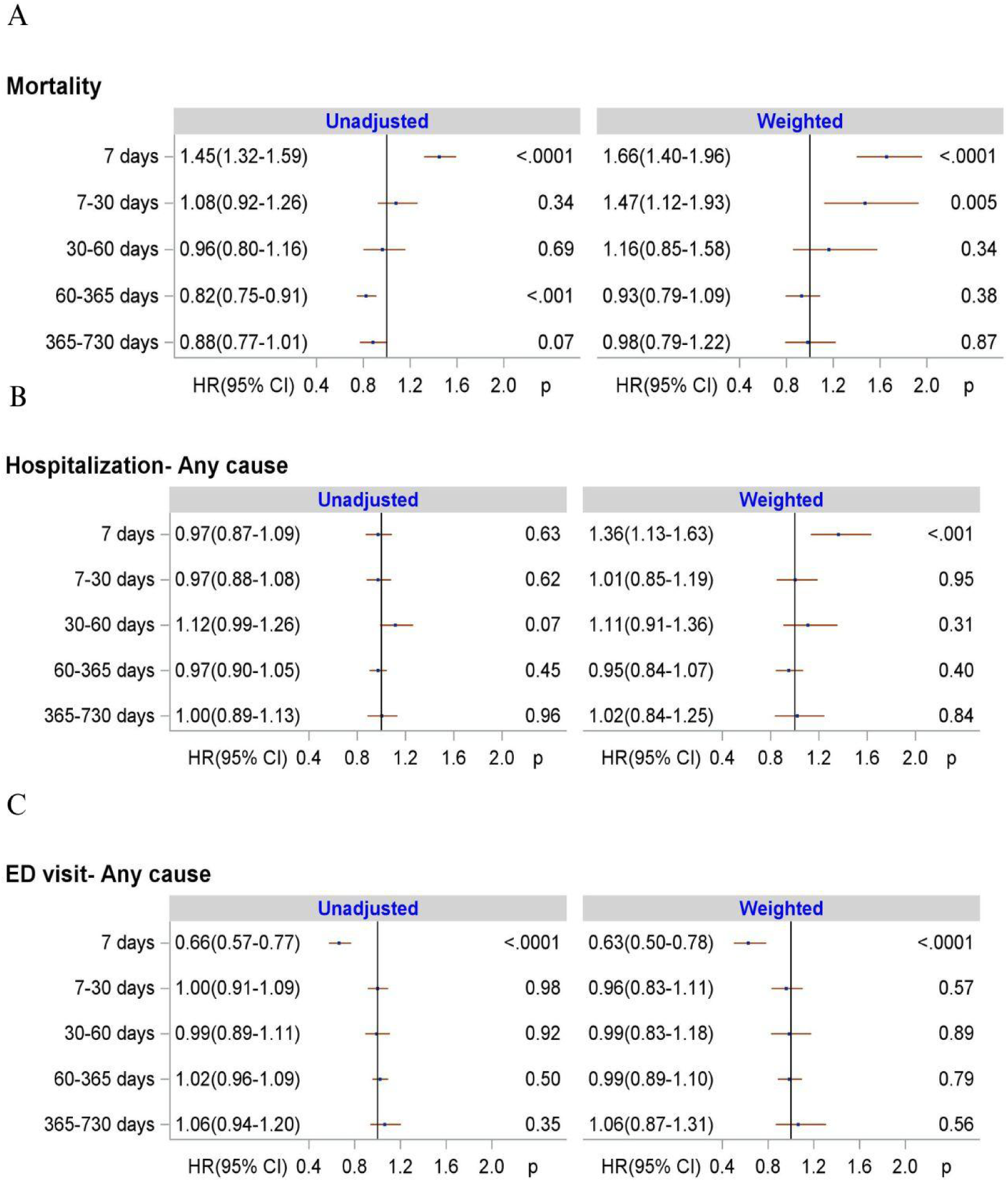
Forest plots showing the unadjusted and weighted hazard ratios (HR) at various time points for risk of any cause mortality (A), any cause hospitalization (B), and any cause emergency department visits (C) for patients who received IV magnesium compared to patients who did not. Abbreviations: ED, emergency department; HR, hazard ratio; CI, confidence interval.

**Figure 3.**
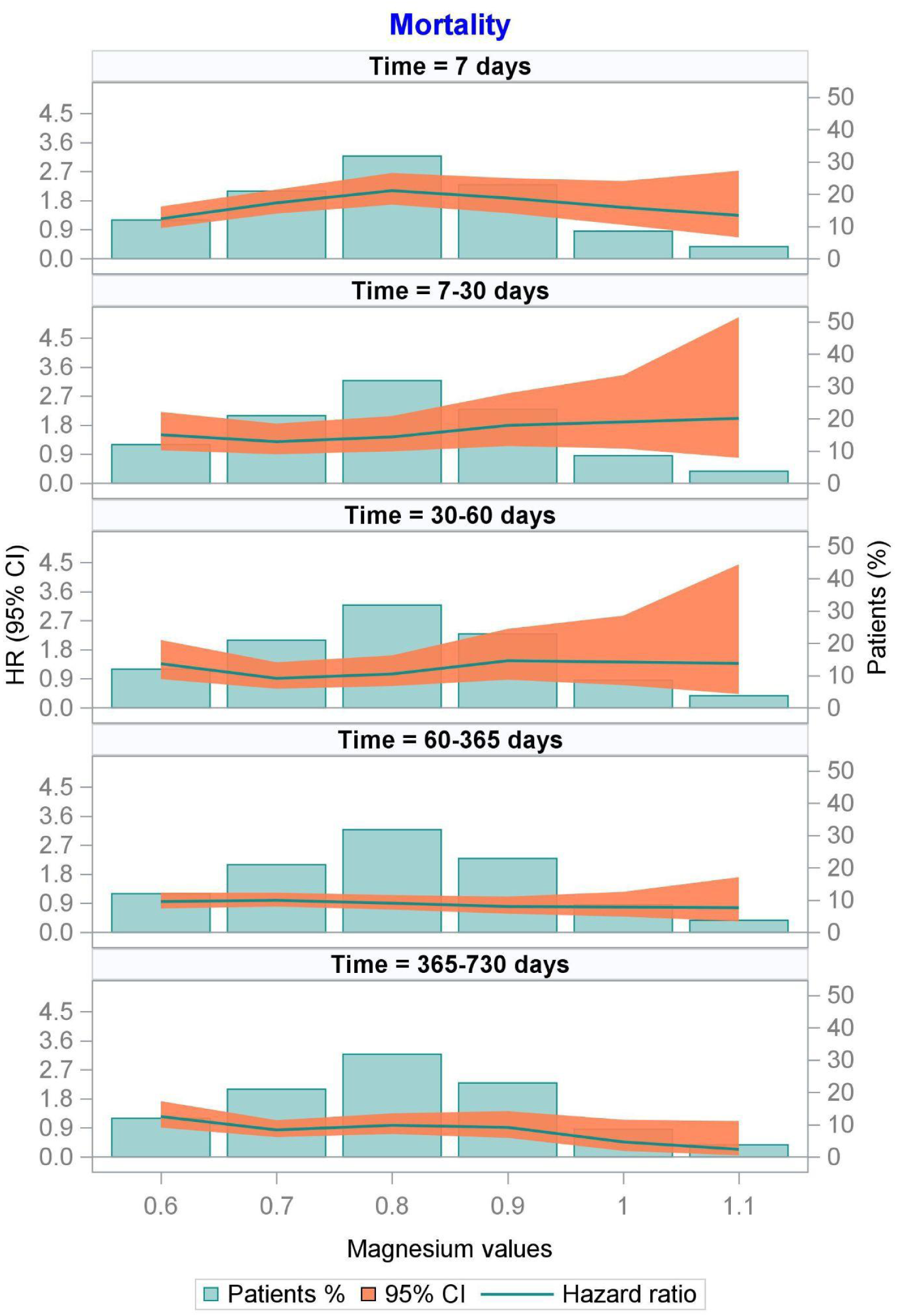
Serum magnesium concentrations of patients who received IV magnesium overlaid with a hazard ratio curve for mortality at various time points. Abbreviations: HR, hazard ratio; CI, confidence interval.

IV magnesium therapy was not associated with risk of hospitalization due to urinary tract infection (UTIs) (HR 2.61 (95% CI 0.68-9.97); p=0.1604) at 7 days or at any other time point, before or after adjustment with propensity weighting **(Supplemental Table 4)**. Neither was IV magnesium therapy associated with risk of hospitalization due to hip fractures at any time point **(Supplemental Table 4)**.

## Discussion

This study explored the factors and outcomes related to serum magnesium testing, hypomagnesemia and IV magnesium supplementation in patients with AHF, which to the best of our knowledge, have not been described before. There are three key results from this study which warrant discussion. First, serum magnesium testing is associated with more medical testing, longer hospital stays and worse patient comorbidity profiles. Second, serum magnesium levels are associated with increased mortality below 0.7 mmol/L and hypomagnesemia is not associated with higher BNP or NT-proBNP levels - not previously examined. Finally, and contrary to presumed safety or effectiveness, IV magnesium administration was associated with greater short-term mortality. While not definite, this evidence should raise concern to the clinical community and warrants investigation through pragmatic clinical trials.

We determined that serum magnesium testing occurs frequently for patients with HF and that it is more likely with longer hospital stays, and with tertiary care hospitals. Patients who were tested for serum magnesium were also more likely to have been tested for other markers and electrolytes such as sodium, potassium and creatinine. This suggests that testing was more likely to occur in episodes within bulk testing rather than targeted laboratory assessment due to the complexity of patient conditions or competing diagnoses. Additionally, the likelihood of receiving IV magnesium supplementation was high for patients with AHF and was even higher when patients were hypomagnesemic, were admitted rather than discharged and when patients visited tertiary hospitals.

There is no consensus concerning the prognostic significance of hypomagnesemia in HF and limited agreement concerning the significance or factors related to hypomagnesemia in AHF. Our study found that serum magnesium concentrations were associated with mortality when they were below the level of 0.7 mmol/L and above the level of 0.86 mmol/L. In comparison to studies in the past which used various a priori cutoffs to define hypomagnesemia and have found both evidence for and against an association between low magnesium levels and poor outcomes, our study evaluated magnesium as a continuous variable;^3–11, 15, 17^ For example, the EVEREST study, which was the only other study to evaluate the prognostic significance of serum magnesium in AHF, used a similar cutoff of 0.74 mmol/L to our data driven cut off of 0.7 mmol/L for the hypomagnesemic group, however the mean magnesium concentration in this group was 0.7 mmol/L which suggests that much of this group may not have had magnesium levels which would be associated with worse future outcomes according to the current results.^11^

While our study provides more evidence supporting the prognostic significance of magnesium in AHF, it is still unclear whether serum magnesium is a clinically relevant marker of magnesium related cardiovascular pathophysiology as hypomagnesemia may be associated with poor outcomes for other reasons (eg. lack of high magnesium foods).^5, 14,18–21^ It is also clear that testing is often completed without an intervention in mind and abnormal test results may not change management plans as only 30.3% of patients with hypomagnesemia received IV magnesium. Consequently a greater understanding of the relationship between serum magnesium concentration and pathophysiological processes as well as the patterns of magnesium level testing in AHF are necessary to develop evidence based recommendations about when to test patients and what test results should change patient management.

The associations between IV magnesium administration and hypomagnesemia as well as the Charlson Comorbidity Index and comorbidities such as atrial fibrillation in our study support that magnesium administration occurred in two main patterns: to correct hypomagnesemia as a lab value and in patients with a presumptive heightened risk of arrhythmias. Prior small randomized controlled trials evaluating the efficacy of IV magnesium in chronic or acute HF have focused on using oral or IV magnesium supplementation to produce positive rhythmic, electrolytic, and hemodynamic effects.^7–9, 22–23^ These trials have shown that IV magnesium infusion in patients with chronic and worsening HF results in fewer arrhythmic episodes and greater vascular function.^7–9, 22^ While IV magnesium may be able to reduce arrhythmic events and improve vascular function, the results of our study suggest novel findings of a strong association between magnesium infusion and greater mortality risks at 7 days and between 7-30 days that persisted after adjustment for known confounders. Prior trials evaluating the outcomes of IV magnesium infusion in patients with acute myocardial infarction found no association between mortality and IV magnesium infusion.^12–13^ No other studies to our knowledge have evaluated the impact of IV magnesium on mortality and morbidity in AHF.

As magnesium is a potent vasodilator and may have a role in modulating the central nervous system, it is possible that IV magnesium administration contributed to hypotension and respiratory depression in some patients.^24^ However, our results also indicate that the risk of mortality associated with IV magnesium infusion was greatest for individuals with serum magnesium concentrations in a relatively normal range, and neutral or near neutral for individuals with hyper- and hypo-magnesemia compared to patients who did not receive IV magnesium. This suggests that the association between IV magnesium and increased risk of mortality is unlikely to be dose related. Confounding due to clinical severity, which may be associated independently with both IV magnesium administration and mortality, is tempered as an explanation for our study results due to the lack of an association between IV magnesium administration and negation outcomes.^25–28^ However, clinical severity is a difficult quality to capture in the context of acute heart failure and as a result it is challenging to evaluate the influence of clinical severity on the association between IV magnesium use and mortality in a retrospective manner, particularly since some clinical details were not available.

Randomized controlled trials are necessary to better understand the efficacy and safety of IV magnesium in AHF while accounting for potential confounders such as clinical severity and evaluating the impact of timing, dosing and clinical characteristics of patients, such as comorbidities and EF, on IV magnesium efficacy. Current guidelines do not address the use of IV magnesium in acute heart failure and this is likely due to the lack of any evidence evaluating its impact on mortality and morbidity, as a result the current use patterns of IV magnesium lack a foundation in clinical evidence and should be reconsidered.^29,30^

### Strengths and Limitations

Our study has a number of strengths, but due to its observational and retrospective design it also has some limitations. By using administrative health data from a single payer universal healthcare system, the study population represents the characteristics of the general population and consequently the results are generalizable. The administrative health databases did not have access to data on some clinical characteristics of patients like blood pressure, New York Heart Association functional class, cardiac troponin levels, ischemic etiology or ejection fraction in the case of some patients, however we were able to access and account for other characteristics including laboratory tests like serum potassium, and natriuretic peptide levels.

We addressed the influence of known confounders and factors influencing exposure risk through a propensity weighted analysis and negation endpoint analysis, but we also recognize that there may be unknown or unmeasured variables which may have impacted results. Furthermore, the retrospective observational nature of the study prevents the establishment of a direct causal relationship between associated variables in our study, consequently the relationship between IV magnesium and higher risk of mortality found in the study should be further evaluated through a more rigorous study design such as a randomized controlled trial.

## Conclusion

While serum magnesium testing, hypomagnesemia and IV magnesium supplementation are clinically prevalent, their use as a treatment in AHF may not provide patient benefit or alter patient management; on the contrary, results indicate there may be a harm associated with this treatment in patients with AHF. It is important to further evaluate the relationship between AHF and magnesium and when or if IV magnesium’s use is efficacious, or safe in patients with AHF through randomized controlled trials.

## Data Availability

All data produced in the present study are available upon reasonable request to the authors.

## Acknowledgements

Data were extracted from the Alberta Health Services Enterprise Data Warehouse with support provided by AbSPORU Data and Research Services platform which is funded by CIHR, Alberta Innovates, University Hospital Foundation, University of Alberta, University of Calgary and Alberta Health Services. The interpretation and conclusions contained herein are those of the researchers and do not necessarily represent the views of Alberta Health Services or any of the funders. The authors would like to gratefully acknowledge Lisa Soulard for editorial assistance in preparing the manuscript.

