## Supplementary Material for "Outcomes and Patterns Related to Magnesium in Acute Heart Failure: A Population-Based Study"

### Supplemental Material

Supplemental Methods……………………………………………………………………page 2-4

Supplemental Table 1. ICD- 9 and 10 codes…………………………………………………page 5

Supplemental Table 2. Number and crude rate (per 100 person-months) for all patient outcomes……………………………………………………………………………page 6

Supplemental Table 3. Factors associated with likelihood of IV magnesium administration………………………………………………………………………page 7-8

Supplemental Table 4. Association between falsification endpoints and IV magnesium administration………………………………………………………………………..page 9

Supplemental Figure 1. Propensity score distribution before (A) and after (B) overlap weighting……………………………………………………………………………..page 10

Supplemental Figure 6. Forest plots with unadjusted and adjusted hazard ratios for other patient outcomes after IV magnesium administration from a time-varying Cox proportional hazard model………………………………………………………………………………page 15-16

**Supplemental Methods**

*Data sources*

Through the discharge abstract database (DAD) and the national ambulatory care reporting system (NACRS) data we were able to identify all primary heart failure hospitalizations or emergency department visits (ED) during the mentioned time period. The Pharmaceutical Information Network (PIN) database provided the information (including Drug Identification Number (DIN), Anatomical Therapeutic Chemical Classification (ATC), dispensing date, amount and days of supply) on prescribed medications dispensed to outpatients in Alberta from January 1, 2008 onwards. Hospital-based pharmaceutical datasets including DOSE and SCM were used to obtain information on IV magnesium supplementation. The laboratory testing details were collected from a province-wide laboratory repository that included inpatient, ED and outpatient lab tests and was available from April 2012 onwards. The Population Registry database contained patients' demographic information (i.e. year of birth, sex, first three digits of patients' residential postal code) and the Vital Statistics database provided information on deaths including date of death.

*Patient Population*

We considered ED visits and hospitalizations with HF within 48 hours as a single episode of care. Patient comorbidities were identified using ICD codes (9th and 10th revision) from any hospitalizations, ambulatory encounters, or from physician's claims in the outpatient setting (≥2 claims at least 30 days apart within one year) for all healthcare encounters up to six years prior to the index episode.

*Intravenous Magnesium Therapy Classification*

Administration of magnesium sulfate was only considered if it occurred within ± 1 day of HF episodes.

*Outcomes*

Patients in the cohort were followed from each episode for two years until they had another episode of HF hospitalization or ED visits, moved out of the province, died or the study ended (March 31, 2020), whichever occurred first. However, the next episode of HF hospitalization or ED visit, that occurred in two years, might be considered as the outcome of the earlier episode.

*Statistical Analysis*

Variables evaluated in the General Estimating Equations and the time varying Cox regression models with weighting included: patient demographics (age and sex), residence, hospital type, clinical setting (ED and admission), fiscal year, diabetes, hypertension, hyperlipidemia, CAD, PVD, Afib, thromboembolism, liver disease, renal disease, asthma, COPD, cancer, depression, dementia, medications, comorbidities and laboratory test results (sodium, potassium, magnesium, serum creatinine and hemoglobin). Linearity assumption for the relationship of continuous variables such as age with magnesium testing and hypomagnesemia was assessed and included as the natural cubic spline including percentiles with 3 knots.

We limited the time-varying Cox regression models with weighting analysis to patients aged 60 to 90 years old, with a maximum of three HF episodes, due to the lower number of IV magnesium administration events among patients who did not meet the above criteria preventing effective group balancing.

Around 99% of patients had hemoglobin and serum creatinine tests available, while 67.5% had sodium and potassium tests available. Missing sodium, potassium, hemoglobin and creatinine values were imputed using multiple imputation with a fully conditional specification method. After examining the interaction between variables, an interaction term for age and serum magnesium value was included in the propensity model. The propensity score was then calculated and included as a time varying covariate and the balance of variables between groups was assessed using the standardized mean differences; variables were considered well balanced if the standardized mean differences ranged from -0.2 to 0.2.

**Supplemental Table 1.** ICD- 9 and 10 codes

| **Comorbidities** | **ICD-10-CA** | **ICD-9** |
|---|---|---|
| Diabetes | E10, E11, E12, E13, E14 | 250 |
| Hypertension | I10, I11, I12, I13, I15 | 401, 402, 403, 404, 405 |
| Dyslipidemia | E785 | 272.4 |
| CAD | I250, I251, I252, I255, I258, I259 | 411, 412, 413, 414 |
| PVD | I70, I71, I731, I738, I739, I771, I790, I792, K551, K558, K559, Z958, Z959 | 093.0, 437.3, 440, 441, 443.1, 443.2, 443.3, 443.4, 443.5, 443.6, 443.7, 443.8, 443.9, 447.1, 557.1, 557.9, V43.4 |
| Atrial Fibrillation | I48 | 427.3 |
| Thromboembolism | I26, I801, I802, I803, I808, I809, I828, I829, I821, I822, I823, I26, I801, I802, I803, I822, I823, I828, I829, I74 | 415, 451.1, 451.8, 451.9, 453.1, 453.2, 453.4, 453.5, 453.8, 453.9, 415, 451.1, 451.2, 453.3, 453.8, 453.9, 444 |
| Asthma | J45 | 493 |
| COPD | J40, J41, J42, J43, J44, J45, J46, J47, J60, J61, J62, J63, J64, J65, J66, J67, I278, I279, J684, J701, J703 | 491, 492, 494, 496 |
| Cancer | C0, C1, C20, C21, C22, C23, C24, C25, C26, C30, C31, C32, C33, C34, C37, C38, C39, C40, C41, C43, C45, C46, C47, C48, C49, C50, C51, C52, C53, C54, C55, C56, C57, C58, C60, C61, C62, C63, C64, C65, C66, C67, C68, C69, C70, C71, C72, C73, C74, C75, C76, C81, C82, C83, C84, C85, C88, C90, C91, C92, C93, C94, C95, C96, C97 | 14, 15, 16, 170, 171, 172, 174, 175, 176, 177, 178, 179, 18, 190, 191, 192, 193, 194, 195, 200, 201, 202, 203, 204, 205, 206, 207, 208, 238.6, 196, 197, 198, 199 |
| Sleep Apnea | G473 | 327.2 |
| Depression | F204, F313, F314, F315, F32, F33, F341, F412, F432 | 296.2, 296.3, 296.5, 300.4, 309, 311 |
| Dementia | F00, F01, F02, F03, F051, G30, G311 | 290, 294.1, 331.2 |
| **Medications** | **ATC** | |
| ACEi | C09A, C09B | |
| ARB | C09C, C09D | |
| ARNi | C09DX04 | |
| MRA | C03DA | |
| Beta blocker | C07 | |
| Digoxin | C01AA05 | |
| Statin | C10AA, C10BA, C10BX | |

**Supplemental Table 2.** Number and crude rate (per 100 person-months) for all patient outcomes.

| Outcomes | No supplement | IV Mg supplement |
|---|---|---|
| **Mortality** | 9010 (2.6) | 1539 (2.7) |
| **Hospitalization** | | |
| CV | 7562 (2.6) | 1214 (2.5) |
| HF | 5141 (1.6) | 762 (1.4) |
| Any cause | 13339 (5.8) | 2176 (5.8) |
| **ED visit** | | |
| CV | 8409 (2.9) | 1369 (2.8) |
| HF | 6218 (1.9) | 1021 (1.9) |
| Any cause | 14974 (7.5) | 2452 (7.2) |
| **Physicians' claim** | | |
| CV | 16334 (13.8) | 2822 (16.4) |
| HF | 12552 (6.9) | 2148 (7.5) |
| Any cause | 18040 (19.4) | 3071 (22.7) |
| **Hospitalization** | | |
| UTI | 544 (0.2) | 85 (0.2) |
| Hip fracture | 188 (0.1) | 25 (0.0) |

Abbreviations: ED, emergency department; CV, cardiovascular; HF, heart failure; UTI, urinary tract infection.

**Supplemental Table 3.** Factors associated with likelihood of IV magnesium administration

| **Variables** | **aOR (95% CI)** | **p-value** |
|---|---|---|
| Age <75 years (per 5 years increase) | 0.97 (0.95-0.99) | 0.003 |
| Age ≥75 years (per 5 years increase) | 0.76 (0.73-0.78) | <0.001 |
| Sex- Males vs Females | 1.18 (1.10-1.26) | <0.001 |
| Residence- Urban vs Rural | 1.55 (1.42-1.69) | <0.001 |
| Hospital- tertiary vs non-tertiary | 1.35 (1.26-1.45) | <0.001 |
| Clinical setting- Admitted vs ED discharged | 2.41 (2.22-2.62) | <0.001 |
| Fiscal year | | <0.001 |
| 2013 vs 2012 | 1.07 (0.93-1.22) | |
| 2014 vs 2012 | 1.23 (1.08-1.40) | |
| 2015 vs 2012 | 1.26 (1.11-1.43) | |
| 2016 vs 2012 | 1.19 (1.05-1.36) | |
| 2017 vs 2012 | 1.28 (1.13-1.46) | |
| 2018 vs 2012 | 1.38 (1.21-1.56) | |
| 2019 vs 2012 | 1.46 (1.26-1.68) | |
| Mg level ≤1.1 (per 0.1 unit decrease) | 1.28 (1.19-1.38) | <0.001 |
| Mg level ≤0.8 (per 0.1 unit decrease) | 4.08 (3.92-4.25) | <0.001 |
| Diabetes | 0.83 (0.77-0.89) | <0.001 |
| Hypertension | 0.89 (0.81-0.98) | 0.024 |
| Hyperlipidemia | 0.93 (0.86-1.01) | 0.091 |
| CAD | 1.13 (1.06-1.21) | <0.001 |

| | | |
|---|---|---|
| PVD | 1.03 (0.94-1.12) | 0.52 |
| Atrial fibrillation | 1.17 (1.09-1.25) | <0.001 |
| Thromboembolism | 0.98 (0.90-1.07) | 0.65 |
| Asthma | 1.00 (0.90-1.10) | 0.96 |
| COPD | 0.85 (0.80-0.92) | <0.001 |
| Anemia | 0.93 (0.86-0.99) | 0.033 |
| Cancer | 1.00 (0.92-1.09) | 0.98 |
| Sleep Apnea | 0.93 (0.84-1.02) | 0.13 |
| Depression | 0.89 (0.83-0.97) | 0.005 |
| Dementia | 0.94 (0.84-1.05) | 0.27 |
| Smoking | 0.95 (0.87-1.04) | 0.26 |
| ACEi/ARB/ARNi | 0.96 (0.89-1.05) | 0.36 |
| MRA | 0.93 (0.87-1.01) | 0.078 |
| Beta blocker | 0.83 (0.77-0.90) | <0.001 |
| Digoxin | 0.91 (0.83-1.01) | 0.075 |
| Statin | 0.97 (0.90-1.04) | 0.37 |

Abbreviations: ED, emergency department; CAD, coronary artery disease; PVD, peripheral vascular disease; COPD, chronic obstructive pulmonary disease; ACEI, angiotensin converting enzyme inhibitor; ARB, angiotensin-receptor blocker; ARNI, angiotensin receptor neprilysin inhibitor; MRA, mineralocorticoid receptor antagonist

**Supplemental Table 4.** Association between falsification endpoints and IV magnesium administration.

| | Unadjusted | | Weighted | |
|---|---|---|---|---|
| | HR (95% CI) | p-value | HR (95% CI) | p-value |
| Hospitalization- UTI | | | | |
| 7 days | 1.79(0.91-3.50) | 0.0892 | 2.61(0.68-9.97) | 0.1604 |
| 7-30 days | 1.07(0.51-2.27) | 0.8546 | 0.76(0.20-2.89) | 0.6873 |
| 30-60 days | 1.33(0.62-2.87) | 0.4591 | 1.28(0.34-4.85) | 0.7197 |
| 60-365 days | 0.81(0.57-1.16) | 0.2588 | 0.84(0.48-1.48) | 0.5563 |
| 365-730 days | 0.84(0.55-1.29) | 0.4249 | 0.54(0.27-1.08) | 0.0819 |
| Hospitalization- hip fracture | | | | |
| 30 days | 0.51(0.07-3.95) | 0.5214 | 1.19(0.03-45.0) | 0.9241 |
| 30-60 days | 0.88(0.26-2.95) | 0.8356 | 0.69(0.10-4.92) | 0.7135 |
| 60-365 days | 0.94(0.53-1.65) | 0.8227 | 1.16(0.46-2.94) | 0.7532 |
| 365-730 days | 0.68(0.31-1.49) | 0.3333 | 0.62(0.20-1.91) | 0.4028 |

Abbreviations: HR, hazard ratio; UTI, urinary tract infection.

**Supplemental Figure 1.** Propensity score distribution before (A) and after (B) overlap weighting

Abbreviations: IV, intravenous; Mg, magnesium.

A

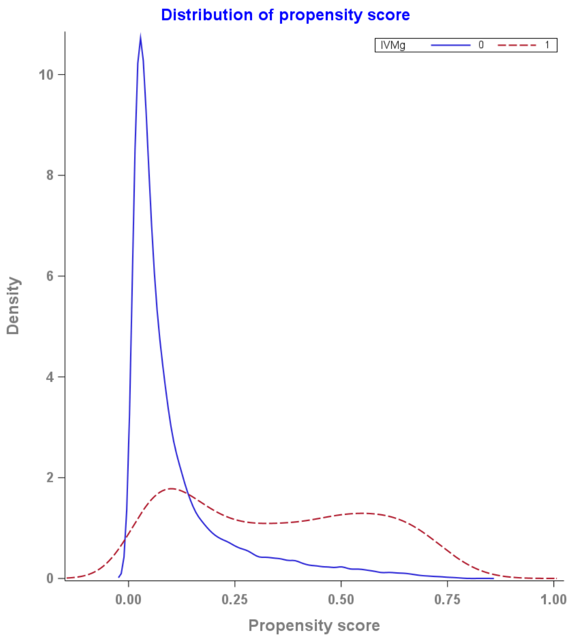

B

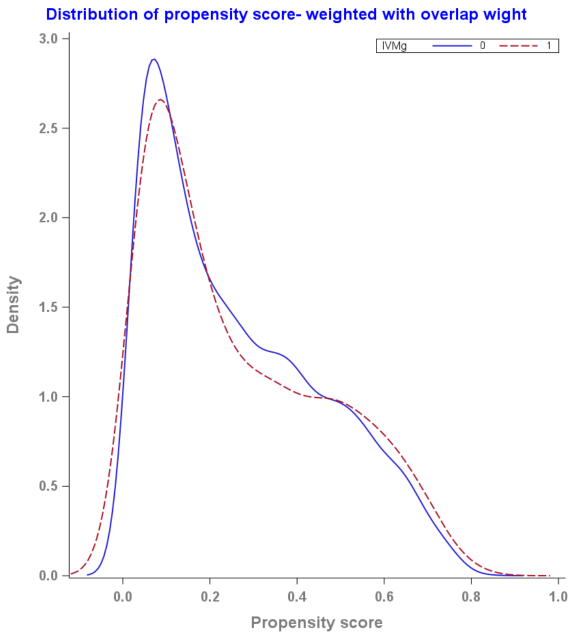

**Supplemental Figure 2.** Forest plot showing the association between demographic and clinical variables and likelihood of serum magnesium testing.

Abbreviations: ED, emergency department; ACEI, angiotensin converting enzyme inhibitor; ARB, angiotensin-receptor blocker; ARNI, angiotensin receptor neprilysin inhibitor.

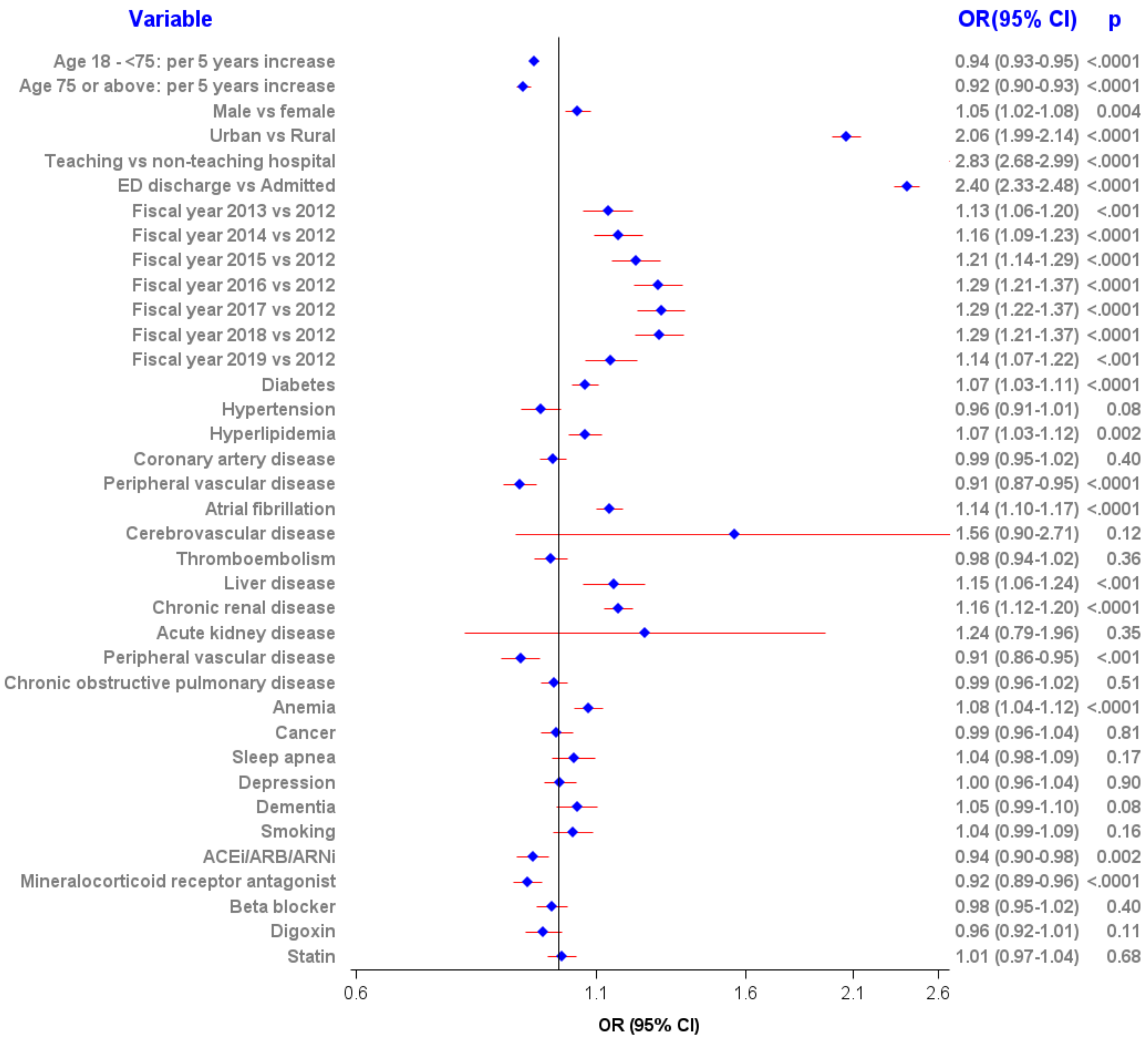

**Supplemental Figure 3.** Forest plot showing the association between demographic and clinical variables and likelihood of hypomagnesemia (<0.65 mmol/L).

Abbreviations: ED, emergency department; ACEI, angiotensin converting enzyme inhibitor; ARB, angiotensin-receptor blocker; ARNI, angiotensin receptor neprilysin inhibitor.

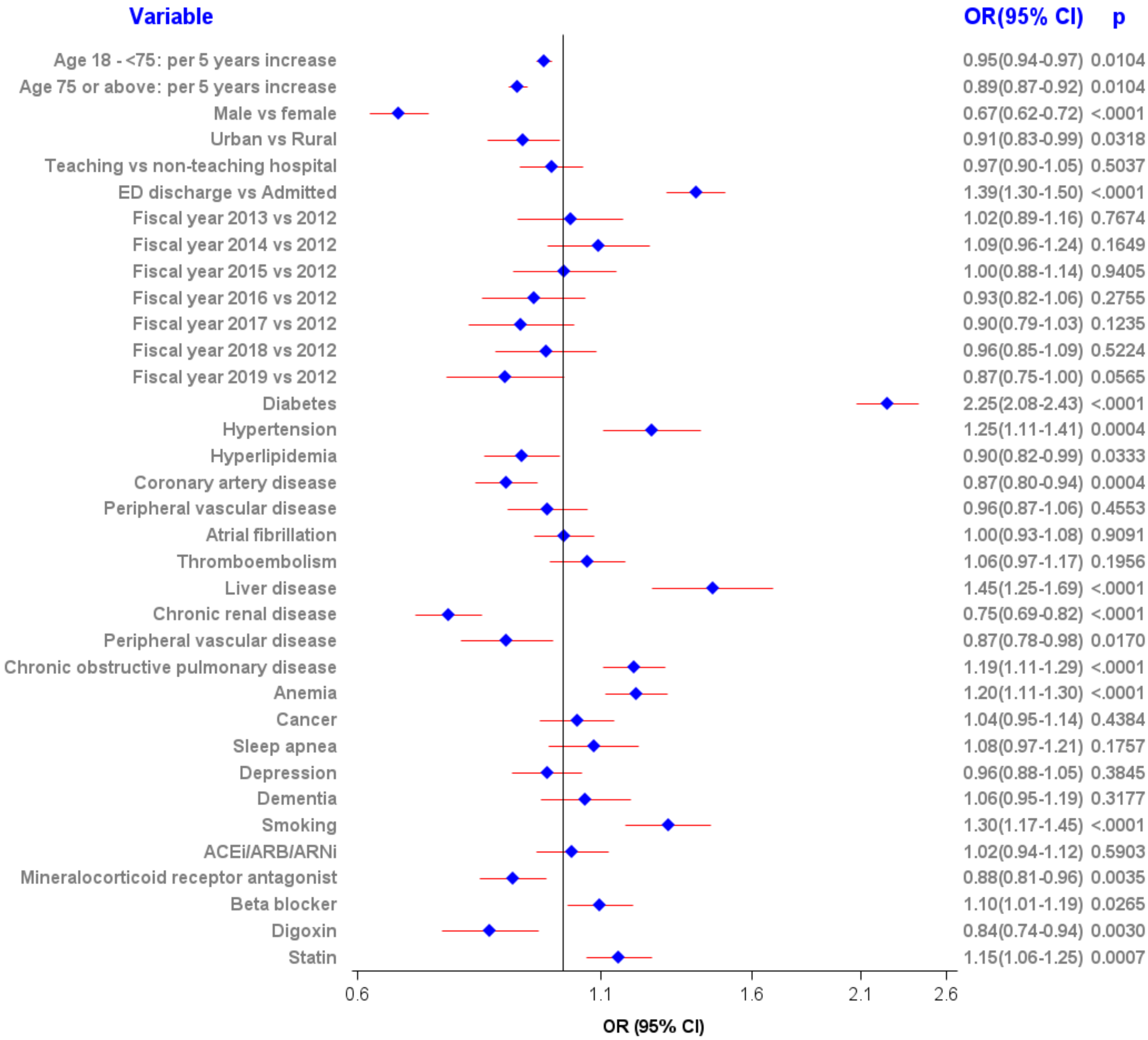

**Supplemental Figure 4.** Forest plot showing the association between demographic and clinical variables and likelihood of hypomagnesemia (<0.70 mmol/L).

Abbreviations: ED, emergency department; ACEI, angiotensin converting enzyme inhibitor; ARB, angiotensin-receptor blocker; ARNI, angiotensin receptor neprilysin inhibitor.

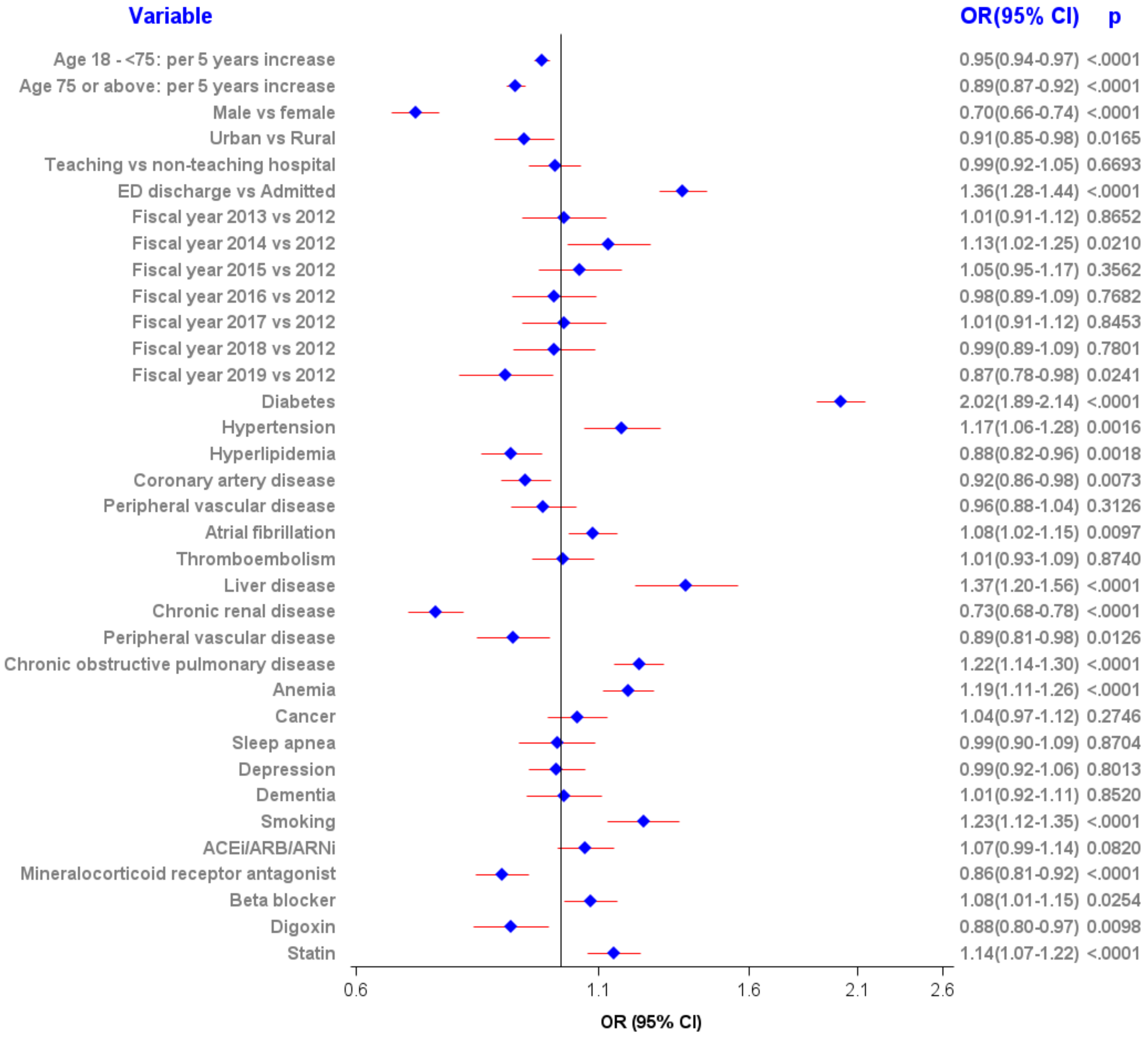

**Supplemental Figure 5.** Forest plot showing the association between demographic and clinical variables and likelihood of hypomagnesemia (<0.75 mmol/L).

Abbreviations: ED, emergency department; ACEI, angiotensin converting enzyme inhibitor; ARB, angiotensin-receptor blocker; ARNI, angiotensin receptor neprilysin inhibitor.

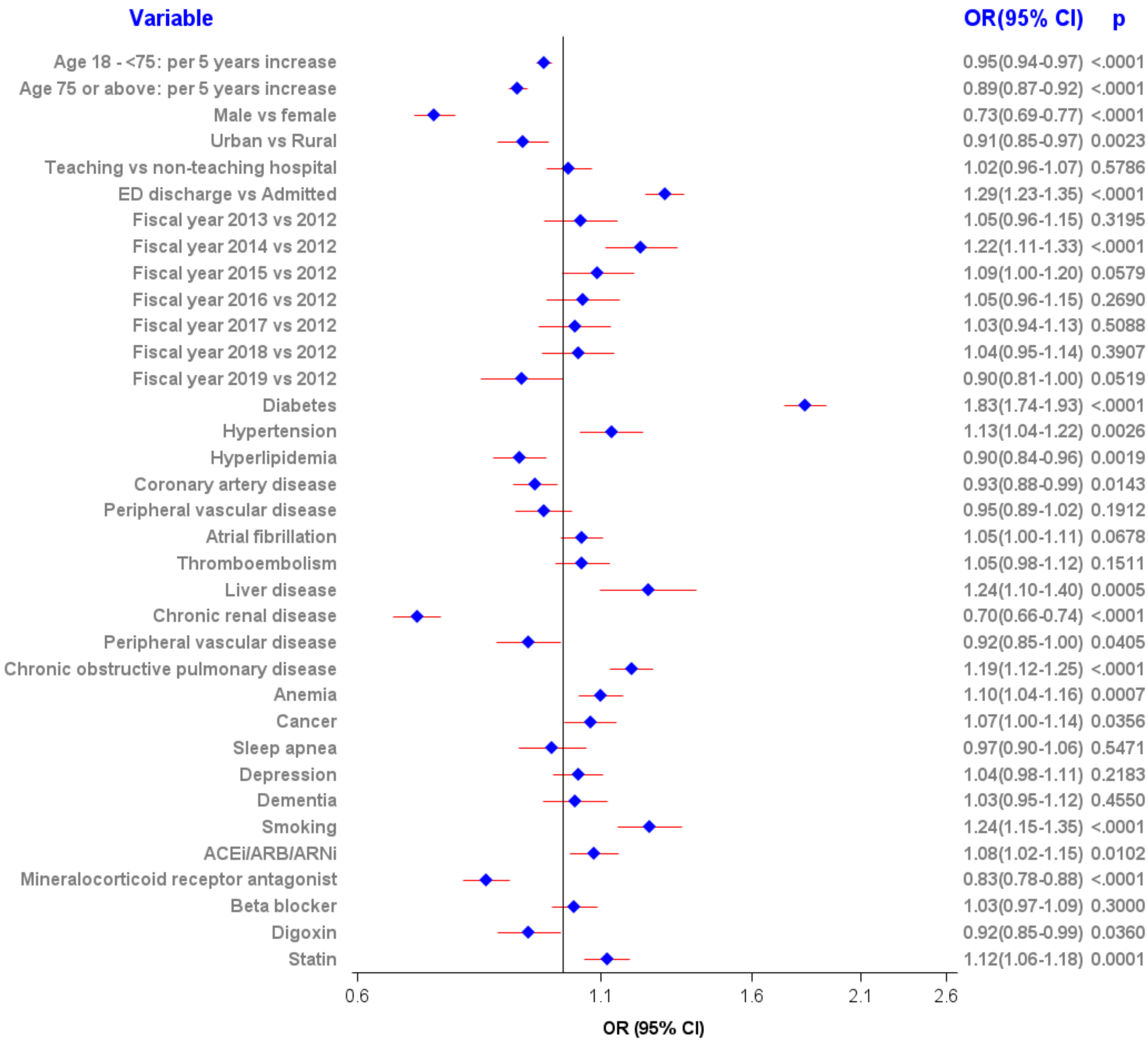

**Supplemental Figure 6.** Forest plots with unadjusted and adjusted hazard ratios for other patient outcomes after IV magnesium administration from a time-varying Cox proportional hazard model.

Abbreviations: ED, emergency department; CV, cardiovascular; HF, heart failure; HR, hazard ratio; CI, confidence interval.

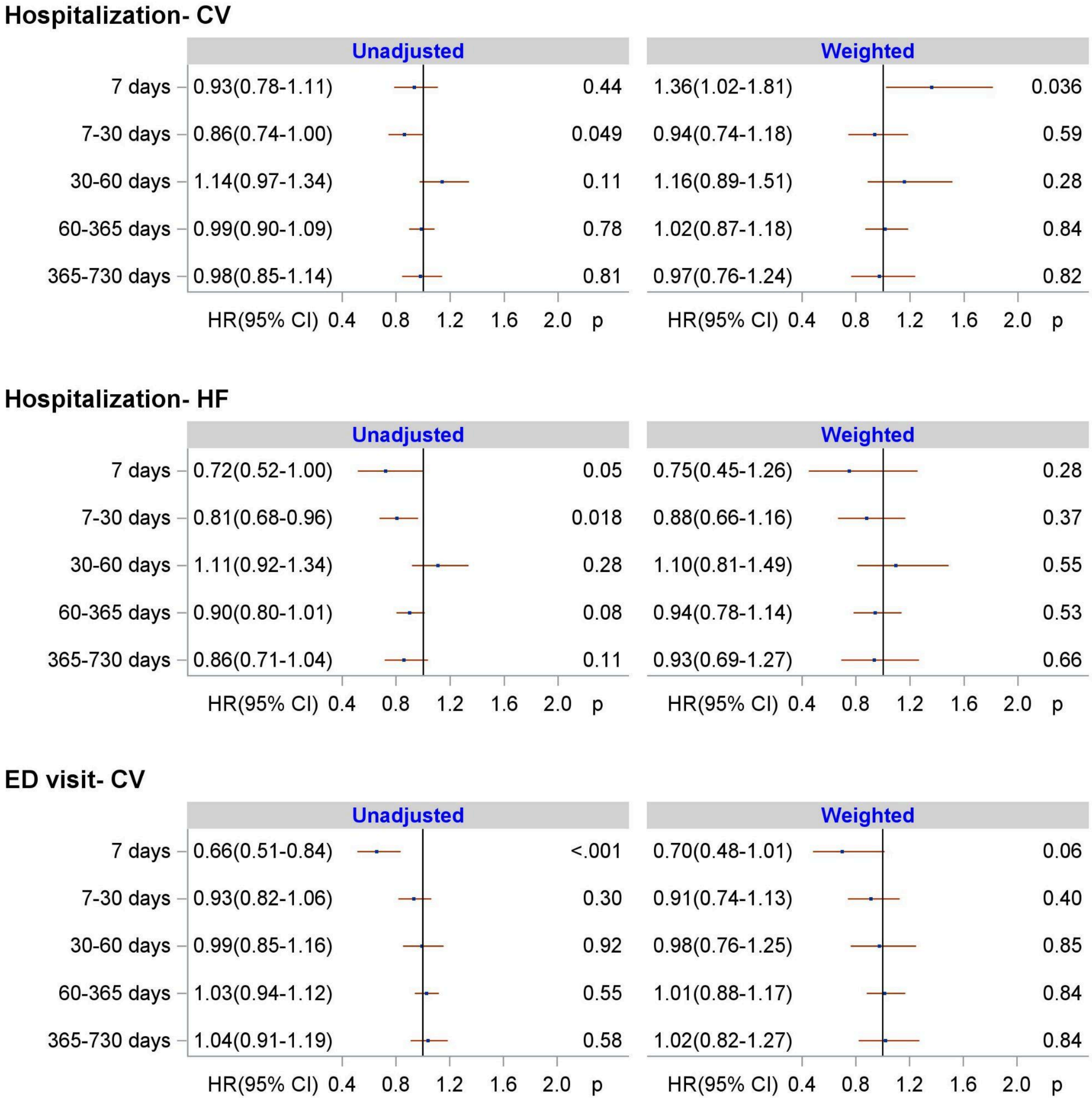

**ED visit- HF**

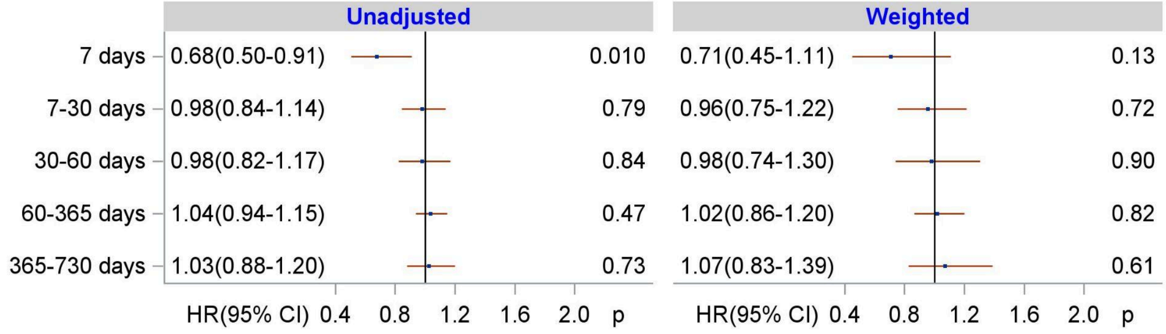

**Physicians claims- Any cause**

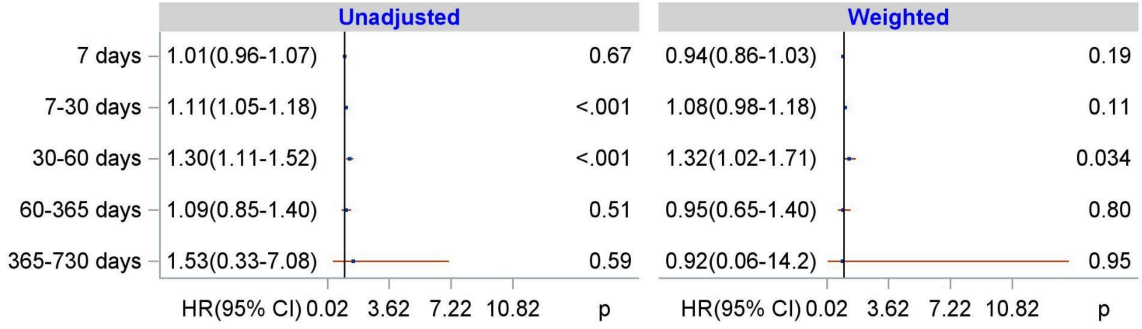

**Physicians claims- CV**

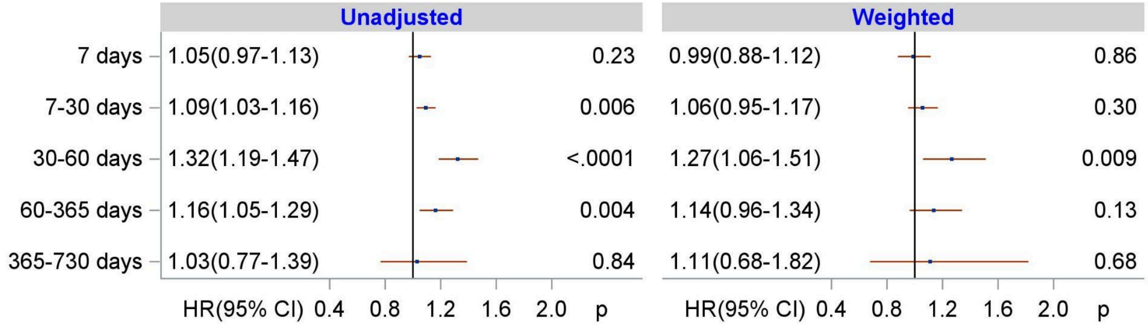

**Physicians claims- HF**

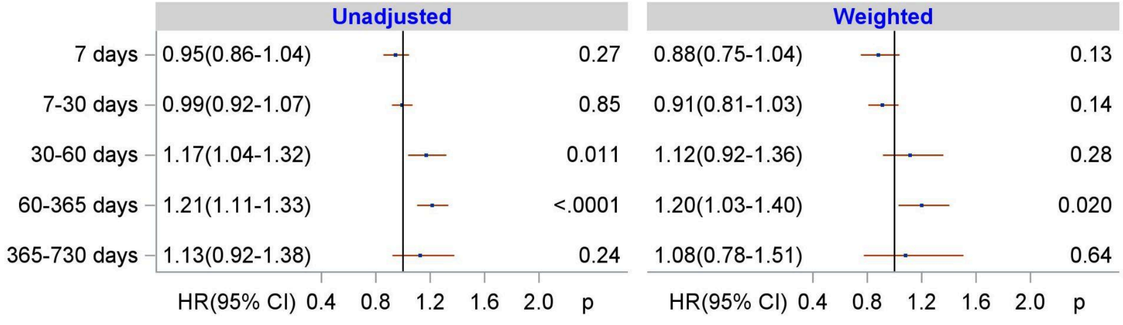
